# Protocol of the observational study STRATUM-OS: First step in the development and validation of the STRATUM tool based on multimodal data processing to assist surgery in patients affected by intra-axial brain tumours

**DOI:** 10.1101/2025.06.12.25329429

**Authors:** Himar Fabelo, Yolanda Ramallo-Fariña, Jesús Morera, Juan F. Piñeiro, Alfonso Lagares, Luis Jimenez-Roldán, Gustav Burström, Miguel A. García, Lidia García-Pérez, Rebeca Falero, Marta González, Sebastián Duque, Consuelo Rodríguez-Jiménez, María Hernández, Julio J. Delgado-Sánchez, Ana B. Paredes, Guillermo Hernández, Pedro Ponce, Raquel León, Jesús M. González-Martín, Francisco Rodríguez-Esparragón, Gustavo M. Callicó, Ana M. Wägner, Bernardino Clavo, the STRATUM Consortium

## Abstract

**Introduction:** Integrated digital diagnostics can support complex surgeries in many anatomic sites, and brain tumour surgery represents one of the most complex cases. Neurosurgeons face several challenges during brain tumour surgeries, such as differentiating critical tissue from brain tumour margins. To overcome these challenges, the STRATUM project will develop a 3D decision support tool for brain surgery guidance and diagnostics based on multimodal data processing, including hyperspectral imaging, integrated as a point-of-care computing tool in neurosurgical workflows. This paper reports the protocol for the development and technical validation of the STRATUM tool.

**Methods and analysis:** This international multicentre, prospective, open, observational cohort study, STRATUM-OS (study: 28 months, pre-recruitment: 2 months, recruitment: 20 months, follow-up: 6 months), with no control group, will collect data from 320 patients undergoing standard neurosurgical procedures to: (1) develop and technically validate the STRATUM tool, and (2) collect the outcome measures for comparing the standard procedure versus the standard procedure plus the use of the STRATUM tool during surgery in a subsequent historically controlled non-randomized clinical trial.

**Ethics and dissemination:** The protocol was approved by the participant Ethics Committees. Results will be disseminated in scientific conferences and peer-reviewed journals.

**Trial registration number:** [Pending Number]

**ARTICLE SUMMARY:** *Strengths and limitations of this study:*

- STRATUM-OS will be the first multicentre prospective observational study to develop and technically validate a 3D decision support tool for brain surgery guidance and diagnostics in real-time based on artificial intelligence and multimodal data processing, including the emerging hyperspectral imaging modality.
- This study encompasses a prospective collection of multimodal pre, intra and postoperative medical data, including innovative imaging modalities, from patients with intra-axial brain tumours.
- This large observational study will act as historical control in a subsequent clinical trial to evaluate a fully-working prototype.
- Although the estimated sample size is deemed adequate for the purpose of the study, the complexity of the clinical context and the type of surgery could potentially lead to under-recruitment and under-representation of less prevalent tumour types.

## INTRODUCTION

Brain and central nervous system cancer was the 12^th^ most common of cancer-related mortality worldwide in 2020 affecting both sexes and all age groups. Notably it was the 2^nd^ leading cause of cancer mortality in young populations (<34 years).^1^ Neurosurgery remains one of the primary treatment modalities for brain tumours, where surgeons aim to achieve a gross total resection of the tumour with the aid of neuronavigation, intraoperative ultrasound, and/or fluorescence guided techniques. Additionally, intraoperative pathology provides a preliminary diagnosis of the tumour type based largely on microscopic analysis. Other adjunctive techniques such as intraoperative neurophysiological monitoring (IONM), help preserve critical brain functions and prevent neurological damage, ultimately improving the patient”s quality of life (QoL).^2^ Despite these advancements, neurosurgeons continue to face several challenges during brain tumour surgeries. These include: i) the difficulty in differentiating tumour margins, since brain tumours (especially gliomas) diffusely infiltrate the surrounding brain tissue;^3^ ii) the brain-shift phenomenon caused by the movement of the brain after performing the craniotomy and starting the resection that affects the accuracy of neuronavigation;^4^ iii) long waiting times for intraoperative pathology consultation that can take up to 45 min, providing a preliminary diagnosis of the tumour type by analysing a small piece of resected tissue; and iv) the lack of tools to ensure complete low-grade tumour resection that has been shown to significantly impact patients” outcome,^5^ (especially in paediatric cases^6^). Moreover, the integration and interpretation of vast amounts of intraoperative data from multiple devices add another layer of complexity to the surgical decision-making process.

To address these challenges, STRATUM^7,8^ aims to develop a 3D decision support tool for brain surgery guidance and diagnostics integrating augmented reality and multimodal data processing powered by artificial intelligence (AI) algorithms. This tool will function as a point-of-care computing system (supplemental **Figure S1.A**) and will be developed using a co-creation methodology that actively involves end-users and other stakeholders. The STRATUM tool will include hyperspectral (HS) imaging (HSI) as an emerging imaging modality in the medical field to enhance intraoperative guidance and diagnosis during the neurosurgical procedures (supplemental **Figure S1.B** and **Figure S2**).^9^ Previous works from several members of the STRATUM consortium in different research projects (HELICoiD,^10^ ITHaCA,^11^ and NEMESIS-3D-CM^12^) have demonstrated, as a proof-of-concept, that this technology is suitable for the intraoperative identification and delineation of brain tumours in real-time.^13–17^ Additionally, the tool is expected to provide a real-time deformation of the magnetic resonance imaging (MRI) within the exposed brain surface for brain-shift compensation during surgery. This will be performed by using advanced mathematical models in combination with the intraoperative multimodal data (HSI, depth information and standard surgical microscope imaging) captured by the STRATUM tool.

The system will be developed and clinically evaluated in three main stages (supplemental **Figure S3**). In Stage 1, a customized multimodal data acquisition system was developed to be used in the observational study of *Stage 2* (STRATUM-OS), which will focused on the multimodal data imaging collection to support the development and technical validation of the STRATUM tool. In *Stage 3* the tool will undergo clinical validation through a subsequent, non-randomized, historically controlled, clinical trial (STRATUM-NRCCT). The historic control in STRATUM-NRCCT will be the subjects recruited in STRATUM-OS. Overall, the STRATUM project aims to: i) optimize the integration and processing of existing and emerging data sources, facilitating timely, efficient and accurate surgical decision-making; ii) maximize tumour resection while minimizing the risk of neurological deficits; iii) reduce anaesthesia duration and related risks; iv) decrease waste associated with repeated pathology analysis; and v) optimize healthcare resource utilization.

The general objective of STRATUM-OS is to collect the necessary data from a cohort of patients affected by intra-axial brain tumours with the standard surgical procedure established in current clinical protocols. STRATUM-OS will pursue the following main objectives:

1. To collect pre-stored and in-situ multimodal data for the development of an intraoperative 3D decision support tool for brain surgery guidance and diagnostics in real-time leveraging AI-based multimodal data processing (STRATUM tool).
2. To technically validate the STRATUM tool, aiming for (1) the intraoperative distinction between tumour and non-tumour areas in the exposed brain surface and (2) the identification of contrast-enhancing tumour (CET) or non-contrast-enhancing tumour (nCET/FLAIR-positive) regions in MRI, through AI-driven processing.
3. To compile a historical control group dataset including patient clinical data, health outcomes, surgical and tumour characteristics, and hospital resource utilization and costs. This dataset will be used in the subsequent non-randomized controlled clinical trial (STRATUM-NRCCT), to assess the safety, effectiveness and cost-effectiveness of the STRATUM tool in brain tumours surgery.

## METHODS AND ANALYSIS

### Study design

STRATUM-OS is an international multicentre, prospective, open, observational cohort study, with a follow-up duration of 6 months, in which the data generated from brain tumour surgeries, including a wide range of intra-axial tumour types, will be collected to meet the objectives of the study. In STRATUM-OS patients will receive standard care as per established clinical protocols, with no modification to their treatment. However, patients will be asked to grant access to their clinical information, complete questionnaires, and provide relevant pre, intra and postoperative information related to the surgical intervention. STRATUM-OS is planned for a duration of 28 months (**Figure 1**) divided in i) a pre-recruitment period of 2 months for the installation of and surgeon training on the acquisition system, ii) a recruitment period of 20 months, and iii) follow-up period of 6 months, including one month for the integration and technical validation of the fully-working STRATUM tool. We anticipate that 320 consecutive patients can be recruited during this study in the 3 clinical sites. The protocol has been drafted in accordance with the Standardised Protocol Items: Recommendations for Observational Studies (SPIROS) statement (supplemental **Table S1**).

**Figure 1.**
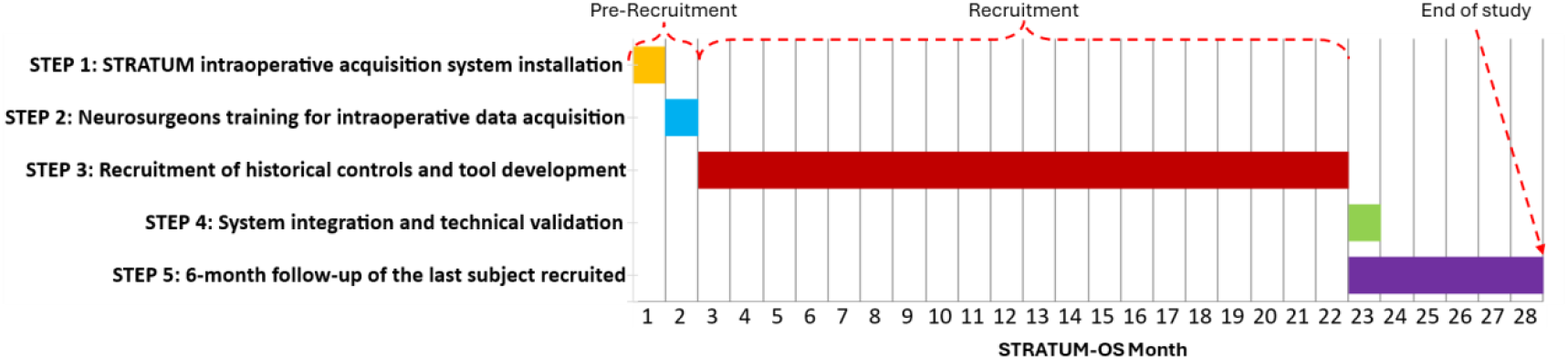
STRATUM-OS Gantt diagram.

### Recruitment and eligibility criteria

Adult participants (≥ 18 years) with an intra-axial brain tumour will be eligible for inclusion. Recruitment will follow a consecutive enrolment process, selecting subjects who meet all the inclusion criteria and none of the exclusion criteria (see **Table 1**) at the participating clinical institutions. Patients will be invited to participate and will be required to sign a written, informed consent form prior to inclusion in the study. They will continue to receive care at their originally assigned medical centre, with no patient transfers between institutions. Members of the research team at each hospital site will introduce the study to subjects who will receive written information describing the study. Researchers will discuss the study details with participants ensuring they have a thorough understanding before making a decision. Participants will have the opportunity to engage in an informed discussion with their physician before consenting. Written informed consent will be obtained from participants or, when applicable, from their designed tutor or legal representatives.

**Table 1:**
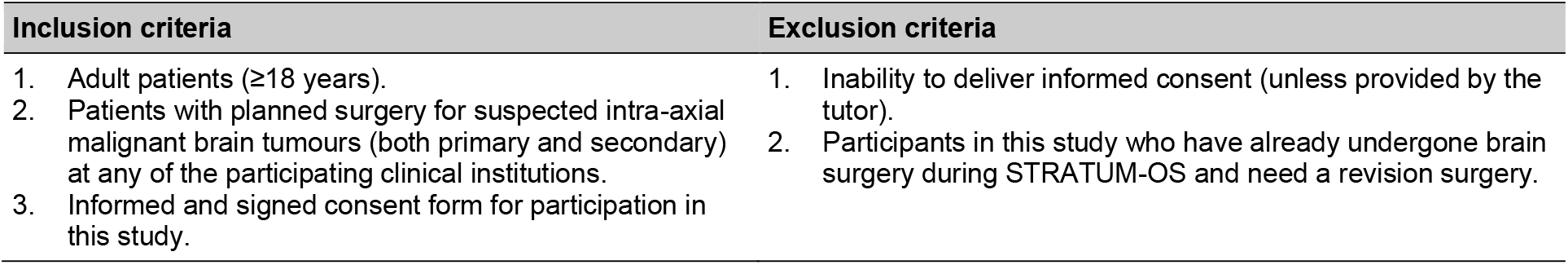
Eligibility criteria for STRATUM-OS.

### Settings

Patients will be recruited at 3 clinical sites: Hospital Universitario de Gran Canaria Doctor Negrín (Las Palmas de Gran Canaria, Spain), Karolinska University Hospital (Solna, Sweden) and Hospital Universitario 12 de Octubre (Madrid, Spain).

### Data collection

The data collection procedure will include data extracted from the EHR of the patient, self-reported questionnaires, information collected from the different professionals involved in the neurosurgical workflow, recorded through an electronic Case Report Form (eCRF), and data collected intraoperatively using the STRATUM acquisition system along with detailed information about the surgery (using the eCRF). The STRATUM eCRF is built on the REDCap (Research Electronic Data Capture) platform^18^ and securely stored in an anonymized and standardized format within a secure repository at the Institute for Applied Microelectronics (IUMA) of the University of Las Palmas de Gran Canaria (ULPGC). REDCap is a secure, web-based software platform designed to support data capture for research studies.^19,20^ It provides an intuitive interface for validated data capture, audit trails for tracking data manipulation and export procedures, automated export procedures for seamless data downloads to common statistical packages and, procedures for data integration and interoperability with external sources. All patient data will be assigned by a unique coded ID [identification] number linked to the subject to ensure pseudo-anonymization. Only the local clinical team will be aware of each participant”s identity. A locally and securely managed document will link each study ID with the corresponding participant. The data collection procedure will be divided in three main phases.

#### Preoperative phase

Patients who meet the inclusion criteria and none of the exclusion criteria and after giving consent to participate in the study will be identified by the Data Collector (DC) at each clinical site. The DC will extract preliminary information from the EHR and confirm the eligibility with the principal investigator at the site before surgery. Preoperative data, including tabular patient information and various preoperative imaging modalities, will be collected, anonymized and transcribed by the DC from several sources (EHR, self-reported questionnaires and interviews/questionnaires/reports from healthcare professionals involved in the neurosurgical workflows). These data will be entered into the STRATUM eCRF.

#### Intraoperative phase

During surgery, the operating surgeon will be assisted by the DC in carrying out the following tasks:

- **Collection of intraoperative data**. The STRATUM acquisition system will be used to capture in-situ HS and standard RGB (Red-Green-Blue) images, depth data, and other relevant intraoperative information of the exposed brain surface.
- **Tumour identification and resection**. The operating surgeon will identify and resect suspicious tumour tissue based on neuronavigation guidance and their surgical judgement according to the standard procedure. At least one tissue sample should be resected from the centre of the tumour site. If possible, the operating surgeon will decide to excise and separately store one to seven additional suspicious tissue samples for definitive pathological diagnosis as part of their routine clinical practice, identifying them within both the STRATUM and the neuronavigation systems for subsequent correlation analysis. Resected tissue samples will be processed according to local protocols at each clinical site. A specific coding system linking the project, clinical site, patient and tissue sample will be used to enter the data in the eCRF. These suspected tumour samples will subsequently undergo histological analysis. Additional samples will be taken from surgical margins where tumour presence is suspected, aligning with the standard surgical procedure. Therefore, no extra sampling of tissue not suspected to be tumorous will be performed.
- **Neuronavigation procedure documentation**. The entire neuronavigation process will be recorded, including multiple positioning points of the neuronavigator marker in relation to the navigable MRI at key stages of the surgical procedure. At least, the neuronavigator marker should be documented within the neuronavigation system (and captured by the STRATUM acquisition system) at the exact tumour site location where the tissue sample will be resected for pathological diagnosis. At least, three HS images will be captured during surgery: 1) A full capture of the exposed brain surface following craniotomy and durotomy; 2) A capture taken at an intermediate stage of the surgery where the tumour tissue is clearly exposed; 3) A final capture after completing the tumour resection. Prior to each HS image, the surgeon will ensure that the exposed area is carefully cleaned to prevent artifacts in the imaging data. When feasible, additional images corresponding to point 2) will be captured throughout the surgery to obtain the clearest possible representation of the brain tumour area.
- **Identification and recording of non-tumour brain tissue data**. At least one highly reliable non-tumour brain tissue area will also be identified and documented based on neuronavigation and the operating surgeon”s judgement. These positions should be recorded within both the neuronavigation system and the STRATUM acquisition system for further correlation analysis, but no tissue sample will be resected. The same coding protocol will be used in the eCRF to label the captured images of non-tumour brain tissue.
- **Intraoperative data storage**. Immediately after surgery, the DC will download the captured data from the STRATUM acquisition system and the neuronavigator and store them in an external, encrypted, hard drive for subsequent pseudo-anonymization. Once pseudo-anonymized, all multimodal data (HS and RGB images, depth data, different MRI modalities, etc.) will be uploaded to the secure, dedicated STRATUM server at IUMA-ULPGC.

#### Postoperative phase

Post-operative data will be collected by the DC from the EHR at one, three, and six months after surgery (see supplemental **Tables S2 to S19**). The same anonymization protocol will be applied, ensuring that all multimodal data is securely stored on the STRATUM server, while tabular data is entered into the STRATUM eCRF.

Once data collection is completed for each patient, the Study Monitor, who is independent of the research team, will review the database for errors and missing data, ensuring data quality. If necessary, the Study Monitor will collaborate with the site DC to resolve errors or discrepancies between the eCRF and the primary source data (e.g., EHR and questionnaires) and make direct revisions in the eCRF. In case of a participant ceases participation in the study or is lost to follow-up, the anonymized data generated until that moment will be employed, if possible, for the technical validation, but the observation will be excluded from the analyses where the missing data is necessary to compute the outcomes in the subsequent clinical trial.

### Outcomes

#### Primary outcome measure for technical validation

The primary outcome measure consists of the histopathological classification of resected tissue samples as “tumour” or “non-tumour”, which will serve as the reference standard for technical validation (supplemental **Table S20**). The STRATUM tool”s performance in intraoperative tumour tissue identification will be evaluated against this reference using a three-way data partitioning approach (training, validation, and test sets), with evaluation metrics including sensitivity, specificity, positive and negative predictive values, and/or receiver operating characteristic (ROC) curve analysis. These performance metrics will be calculated at the tissue sample level using the test set. Assessment will be completed between 3- and 4-weeks post-surgery for the last patient recruited once definitive histopathological results are available.

#### Secondary outcome measures for technical validation

The secondary outcome measure consists of the radiological classification of tumour regions as CET or nCET (FLAIR-positive) based on preoperative and postoperative MRI scans, which will serve as the reference standard for the evaluation of STRATUM”s intraoperative imaging-based segmentation (supplemental **Table S20**). Using a three-way data partitioning approach at the patient level (training, validation and test sets), the agreement between STRATUM-based segmentation and radiological diagnosis will be assessed in the test set using sensitivity, specificity, positive/negative predictive values, Jaccard Index and/or the Dice– Sørensen coefficient. These secondary outcomes will be assessed between 3- and 4-weeks post-surgery for the last patient recruited, following completion and assessment of the post-operative MRI.

#### Data collection for technical validation and the generation of a historical control group

All data collected from patients enrolled in STRATUM-OS will serve as a historical control group for the subsequent study (the non-randomized controlled clinical trial, STRATUM-NRCCT). This trial will compare standard neurosurgical procedures with and without the use of the STRATUM tool for decision-support during surgery. The STRATUM-NRCCT will be conducted upon the completion of STRATUM-OS (see supplemental **Figure S3**). The collected measures will span from patient enrolment to the end of the 6-month follow-up period after surgery and will be categorized in the following main domains (**Table 2**):

**Table 2:**
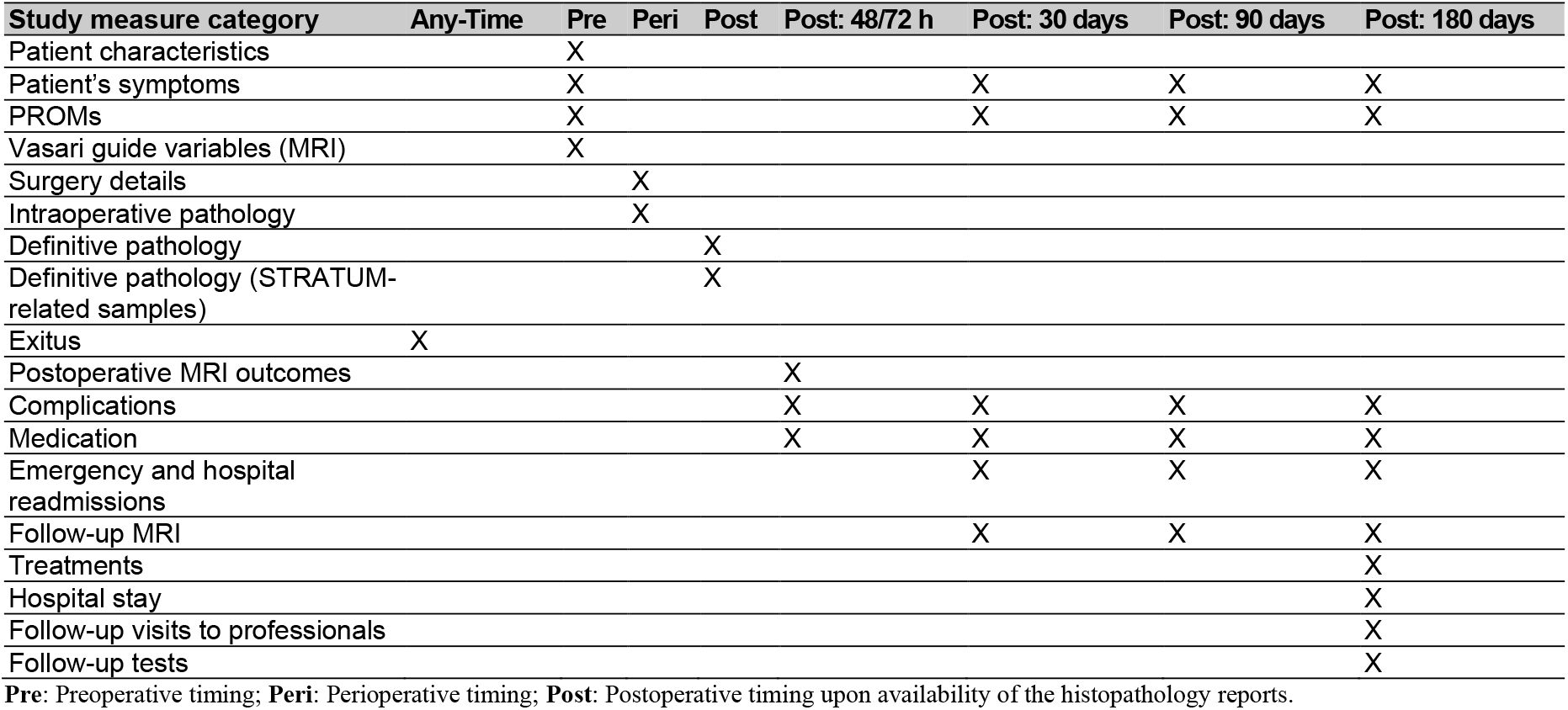
Study measure categories and timing.

- **Patient characteristics**: Tabular data from the patient, such as age, year of birth, weight, sex, etc. (supplemental **Table S2**).
- **Patient’s symptoms**: Tabular data from the patient symptoms related to neurological consequences of the tumour (supplemental **Table S3**).
- **PROMs (Patient-Reported Outcome Measures)**: Scores obtained from the KPS (Karnofsky Performance Status) scale,^21^ the ECOG (Eastern Cooperative Oncology Group) performance status scale,^22^ the 5-level EQ-5D version (EQ-5D-5L) instrument^23^, the EORTC (European Organization for Research and Treatment of Cancer) QLQ-BN20 brain tumour module^24^ and the EORTC QLQ Core Questionnaire (EORTC QLQ-C30)^25^ (supplemental **Table S4**).
- **Vasari guide variables**: The variables related to the radiological characteristics of the brain tumours follow the VASARI (Visually AcceSAble Rembrandt Images) MRI visual feature guide (supplemental **Table S5**).^26^ Such variables will be measured using the minimum requirements to allow lesion volumetry and diagnosis using a 1.5 T MRI.^27,28^
- **Surgery details:** These variables represent the information related to the personnel, tools, materials involved in the surgery, as well as the duration of the different parts and tasks of the surgery (supplemental **Table S6**). These variables will help to quantify the cost and efficiency of the surgery.
- **Intraoperative pathology:** Variables related to the intraoperative tissue diagnosis provided after analysing frozen section samples for rapid diagnosis during surgery according to the local routine practices at each site (supplemental **Table S7**). In case more than one intraoperative tissue analysis is performed, these data will be collected for each sample independently.
- **Definitive pathology**: Variables related to the definitive tissue diagnosis provided by histopathology (supplemental **Table S8**). The CAP (College of American Pathologists) protocol for the examination of tumours of the brain and spinal cord (version 1.0.0.0, September 2022^29^) will be followed to diagnose intra-axial primary brain tumours. Molecular information will be provided according to the 5^th^ edition of the WHO (World Health Organization) Classification of Tumours of the CNS (Central Nervous System),^30^ where it is specified that molecular information should be integrated into many of the tumour types, such as diffuse gliomas and embryonal tumours.
- **Definitive pathology (STRATUM-related samples):** Variables related to the definitive tissue diagnosis provided by histopathology of the additional resected suspected tumour samples for STRATUM technical validation (supplemental **Table S9**). This only includes the specimen size and the tissue type (non-tumour or tumour). The same standard procedures as presented before will be applied to this histopathological analysis.
- **Exitus:** Variables related to the patient”s death in case it occurs (supplemental **Table S10**).
- **Postoperative MRI outcomes**: Variables related to the extent of resection and volume of residual tumour on postoperative 48/72 h MRI using 1.5T MRI (supplemental **Table S11**).
- **Complications**: Set of variables representing the possible postoperative complications related to the brain tumour surgery, including the type of complication, its treatment, and the diagnostic tests performed, if necessary (supplemental **Table S12**).
- **Medication**: Set of variables representing the group type of the medication (painkillers, steroids, antiseizures, other) administered to the patient after brain surgery and related to it (supplemental **Table S13**).
- **Emergency and hospital readmissions**: Variables representing the hospital admission and discharge dates of the patient, including the visits to emergency room, due to causes related to brain tumour surgery, but different to the hospital stay due to the surgical operation (supplemental **Table S14**).
- **Follow-up MRI**: Variables indicating the date of the follow-up MRI, the reason and whether or not progression has occurred (supplemental **Table S15**).
- **Treatments**: This group of variables represents the class of postoperative treatment (chemotherapy or radiotherapy) applied to the patient related to the brain surgery, including the types and subtypes of treatment, the number of cycles or sessions and the dose (in case of chemotherapy) (supplemental **Table S16**).
- **Hospital stay**: Dates of admission and discharge measured from the moment the patient enters (prior surgery) and leaves (after surgery) the hospital, including those related to the time spent at neurosurgical care ward (supplemental **Table S17**).
- **Follow-up visits to professionals**: Variables representing the number of follow-up visits to different healthcare professionals in relation to the brain tumour surgery, including the professional type and the date of the visit (supplemental **Table S18**).
- **Follow-up tests**: Variables representing the number of follow-up tests performed to the patient in relation to the management of the brain tumour, including the type of test and date performed (supplemental **Table S19**).

### Sample size

In total, it is estimated that 26 patients will undergo brain surgery per month across the three clinical sites. Of these, approximately 70% of patients (~18 patients/month) are expected to meet the inclusion criteria, satisfy none of the exclusion criteria, and provide the informed consent for study participation. A 90% success rate in obtaining usable samples is anticipated (~16 patients/month). Consequently, STRATUM-OS is expected to collect data from 320 consecutive patients over 20 months of recruitment, followed by a 6-month follow-up period (total duration: 28 months). Given an average of 4.5 samples per patient (accounting for potential sample loss during pathological analysis), a total of approximately 1,440 tissue samples are expected to be collected.

These estimations have been obtained based on previous experiences from the project partners. Particularly, from the HELICoiD^10^ and ITHaCA^11^ projects in which the same HS acquisition system was employed, a total of 85 HS images were obtained from 41 different subjects captured in three data acquisition campaigns at the Hospital Universitario de Gran Canaria Dr. Negrín, covering a 24-months period in total. From this dataset, 28% of HS images were excluded (17% of subjects), resulting in 61 HS images from 34 eligible subjects.^14^ Additionally, the study conducted at the Hospital Universitario 12 de Octubre within the NEMESIS-3D-CM^12^ project was able to obtain a multimodal dataset composed by HS images from 193 different subjects, also in a 24-months period.^31^

Although patient-level data are important for clinical context and for using them as historical controls in the subsequent STRATUM-NRCCT study, in this study the primary unit of analysis for the main outcome measure is the individual tissue sample, which will be histologically classified as “tumour” or “non-tumour” and serve as the reference standard for technical validation. Therefore, the effective sample size for the statistical analysis is determined by the number of validated tissue samples. This volume of data is expected to provide sufficient statistical power to estimate key diagnostic performance metrics of the STRATUM Tool—such as sensitivity, specificity, and predictive values—with acceptable precision.

A preliminary evaluation of inclusion rates will be conducted after the first 3 months of recruitment, to identify potential barriers to enrolment. If necessary, corrective measures will be implemented to ensure that the study reaches its target sample size.

#### Data partition for technical validation

In AI-based applications, data partitioning is the process of dividing a dataset into several subsets (e.g., training, validation and test sets). This process is crucial to evaluate and validate the performance of developed AI models, ensuring that models are validated and tested using unseen data for model training. This is highly important especially in medical applications, where data from different subjects must be in independent sets.^32^ This allows a more accurate assessment of the model performance, avoiding overfitting and obtaining more generalized models for unseen data/subjects.

In this study, we plan to utilize the initial 70% of recruited patients to train the AI algorithm (*n* = 224 *patients*). The subsequent 10% will be used for cross-validation (*n* = 32), and the final 20% will serve to test the model (*n* = 64). While data partitioning is performed at the patient level to avoid information leakage, the actual analysis will be conducted at the tissue sample level, using labels validated by histopathology to assess diagnostic performance at the tissue sample level.

#### Statistical methods

To ensure a robust and unbiased evaluation of the STRATUM Tool, the dataset will be partitioned at the patient level into three non-overlapping subsets: 70% for model training, 10% for internal validation, and 20% for final testing. This partitioning strategy is intended to prevent data leakage across subsets, ensuring that all tissue samples from the same patient are assigned to the same group. Observations from participants with missing data may be excluded from analyses.

The training set will be used to build the AI model by learning patterns from intraoperative imaging data paired with corresponding histopathological or radiological labels. The validation set will be used during model development to fine-tune hyperparameters and optimize training procedures (e.g., early stopping, learning rate adjustments, regularization). No metrics will be formally reported from the validation set.

The test set will be completely isolated during model development and used exclusively to compute final performance metrics. These will include accuracy, sensitivity, specificity, precision, F1-score, ROC curve analysis, the Jaccard Index, and the Dice– Sørensen coefficient, as appropriate to the outcome type (classification or segmentation). Diagnostic performance will be assessed for two main tasks: (1) the distinction between tumour and non-tumour tissue samples, using definitive histopathological analysis as the reference standard; and (2) the identification of CET and nCET (FLAIR-positive) regions in MRI, using histopathological and radiological reports as the reference.

Subgroup analyses will be performed for the most common histological tumour types, defined as those with at least 25 patients included in the test set. All subgroup evaluations will be conducted exclusively within the test set to ensure unbiased estimation. Due to the subsample nature of the investigation, performance estimates may have wide confidence intervals, particularly for less frequent tumour subtypes.

## DISCUSSION

This protocol describes STRATUM-OS, an international multicentre, prospective, observational cohort study with a 6-month follow-up, designed to collect data from brain tumour surgeries across a range of intra-axial tumour types. The primary aims of STRATUM-OS are to develop and technically validate the STRATUM tool, and to establish a historical control group for the subsequent non-randomized controlled clinical trial (STRATUM-NRCCT). This study addresses critical unmet needs in neurosurgical oncology, such as challenges in tumour margin delineation, brain shift, and the integration of complex intraoperative data. By leveraging multimodal data, including HS imaging and AI-driven analysis, STRATUM-OS has the potential to significantly enhance intraoperative decision-making and improve surgical outcomes. This approach will enable a robust evaluation of the STRATUM tool”s efficacy by comparing outcomes against a baseline derived from real-world clinical practice. Both STRATUM-OS and the subsequent STRATUM-NRCCT contribute to the overarching STRATUM project goal: to advance personalized medicine in neurosurgical oncology through multimodal data, including HS imaging, and AI algorithms, ultimately enhancing intraoperative decision-making, surgical outcomes, and workflow efficiency. In STRATUM-OS, we will collect a comprehensive multimodal dataset from 320 patients undergoing standard neurosurgical procedures for intra-axial brain tumours. For the development and technical validation, the initial 70% of recruited patients will train the AI algorithm, the subsequent 10% will be used for cross-validation, and the final 20% will test the model. To avoid information leakage, data partitioning will occur at the patient level, while the actual analysis will be conducted at the tissue sample level. All patients” data will serve as historical controls for STRATUM-NRCCT. A key strength of STRATUM-OS lies in its prospective, multicentre design, which enhances the generalizability of the findings. Notably, it is the first multicentre prospective observational study to develop and technically validate a real-time 3D decision support tool for brain surgery guidance and diagnostics, integrating AI and multimodal data processing, including HS imaging. This innovative approach, combined with the large sample size, will provide sufficient statistical power for the technical validation of the STRATUM tool. However, the study also has potential limitations, including the complexity of brain tumour surgery, which may slow recruitment and affect sample size, and the potential limitation in subgroup analyses for less common brain tumour subtypes. The rigorous methodology of STRATUM-OS, encompassing detailed data collection protocols, quality control measures, and a comprehensive statistical analysis plan, will ensure the reliability and validity of the results. Furthermore, the study”s adherence to ethical principles and data protection regulations strengthens its design.

## Data Availability

All data produced in the present work are contained in the manuscript

## ETHICS AND DISSEMINATION

The study will adhere to the ethical principles for medical research involving human subjects established in the Declaration of Helsinki and the Good Clinical Practice Guidelines. According to our previous experience (HELICoiD,^10^ ITHaCA,^11^ NEMESIS-3D^12^ and ASTONISH^35^ projects), the use of HSI has not demonstrated any safety or tolerability concerns in surgical procedures. Multimodal data will be captured by expert neurosurgeons using the STRATUM acquisition system designed, produced, and installed at each clinical site (at the time of surgery, no real-time results on tissue classification will be displayed to physicians, except for the standard frozen section histopathological diagnostic information they usually receive). The STRATUM acquisition system will not alter the surgical procedure, apart for the data collection process (estimated to be ~10 min during the entire surgery with no expected negative effects for the patient). Captured data will not influence or modify the neurosurgical plan. As part of the standard procedure, tumour tissue samples and adjacent tissue samples (suspected to be tumour) will be collected for pathological analysis. These samples will serve as golden standard for the algorithm development. This sample collection will not interfere with the intervention, histopathological analysis, or the intraoperative decision-making. Pathologists will have no access to the STRATUM tool results prior to their independent analysis, even during validation and testing phases after initial training.

Patient confidentiality and data security will be managed in compliance with General Data Protection Regulations and relevant national and European legislations as per local and national ethical approvals. All study-related information will be securely stored at the clinical sites. All local databases will be protected by password restricted access systems. Forms, lists, logbooks, appointment books, and any other listings that link participant ID numbers to other identifying information will be stored in a separated, locked restricted-access areas. Only authorized personnel, including researchers involved in the STRATUM project, the sponsor or designated representatives, the Ethics Committee, and relevant health authorities will have access to this data.

All data and biological samples collected during STRATUM-OS will be used exclusively for the development and technical validation of the STRATUM tool, as well as for the creation of a historical control group for the subsequent non-randomized controlled clinical trial (STRATUM-NRCCT). This purpose is explicitly stated among the primary objectives of the present protocol. Therefore, no additional informed consent will be required for this use, as participants will be fully informed and provide consent to both components—technical validation and historical control generation—at the time of inclusion. Any future use of data or samples beyond the scope of this protocol will require prior approval from the relevant Ethics Committees and, where applicable, new participant consent. Participants will have the right to access, rectify, delete, limit the processing, portability and opposition of their data by contacting the principal investigator of the project in each clinical site.

The results of this study will be published in open access journals, regardless of whether the findings are positive or negative. The study results will be shared with the participating physicians, referring clinicians, patients, and the broader medical and scientific community. Data (properly anonymized) will be stored in the secure STRATUM repository at the project coordination institution and, upon project completion, will be archived in trusted repositories, having their respective digital object identifiers (DOIs).

## ACKNOWLEDGEMENTS

Members of the STRATUM Consortium are detailed in https://www.stratum-project.eu/stratum-consortium-members/ and 10.5281/zenodo.15102360. We thank Agustín Quintana Alfonso and Enrique Montesdeoca Ojeda for their invaluable technical support during installation, maintenance, and support of the REDCap platform for this project. The sponsor of the study is the Fundación Canaria Instituto de Investigación Sanitaria de Canarias (FIISC).

## AUTHOR CONTRIBUTIONS

Authorship have followed the criteria set by the International Committee of Medical Journal Editors (ICMJE). All contributors who meet these criteria have been listed as authors. No professional writers or AI-assisted technologies have been used for writing or review the manuscript.

## DISCLAIMER

The funder (European Union) plays no role in design and management of the study, collection, analysis and interpretation of data, writing of the report or the decision on submitting the report for publication.

## PATIENT CONSENT FOR PUBLICATION

Not applicable.

## ETHICS APPROVAL

The Ethics Committee of Hospital Universitario de Gran Canaria Dr. Negrin (Spain), Hospital Universitario 12 de Octubre (Spain) and Karolinska University Hospital (Sweden) have approved this study (No. 2024-395-1 and 25/088), and the study protocol has been registered in ClinicalTrials.gov ([Pending Number]). An insurance policy has been issued for this project under the number 08057767-30092. Any important protocol modifications will be submitted to the relevant Ethics Committees and updated in the trial registry. Investigators will be notified promptly.

## DATA AVAILABILITY STATEMENT

Not applicable.

