## Supplemental Material for "Protocol of the observational study STRATUM-OS: First step in the development and validation of the STRATUM tool based on multimodal data processing to assist surgery in patients affected by intra-axial brain tumours"

### LIST OF SUPPLEMENTAL FIGURES

**Figure S1.** **STRATUM project overall concept**. **A)** Graphical concept of the STRATUM 3D decision support tool, including three main parts: an acquisition system, a processing platform and a graphical user interface. Guidance maps are generated by using an emerging imaging modality, called hyperspectral imaging (HSI), to identify the tumour tissue boundaries in real-time during the surgical procedure. **B)** Traditional imaging versus HSI concept, where HSI provides a vector of reflectance values for each pixel called spectral signature, with a unique pattern for each material/substance/tissue. **RGB**: Red-Green-Blue; **HPC**: High Performance Computing; **PET**: Positron Emission Tomography; **CT**: Computed Tomography; **AR**: Augmented Reality; **GUI**: Graphical User Interface. .......... 4

### SUPPLEMENTAL FIGURES

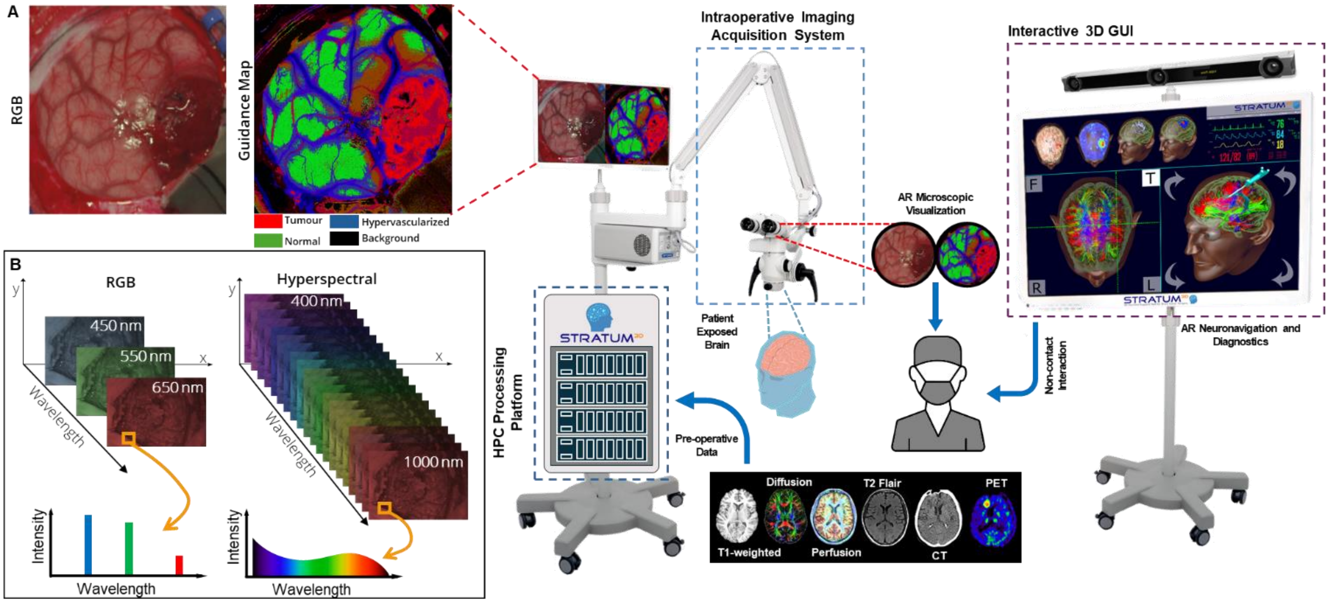

**Figure S1.** STRATUM project overall concept. A) Graphical concept of the STRATUM 3D decision support tool, including three main parts: an acquisition system, a processing platform and a graphical user interface. Guidance maps are generated by using an emerging imaging modality, called hyperspectral imaging (HSI), to identify the tumour tissue boundaries in real-time during the surgical procedure. B) Traditional imaging versus HSI concept, where HSI provides a vector of reflectance values for each pixel called spectral signature, with a unique pattern for each material/substance/tissue. **RGB**: Red-Green-Blue; **HPC**: High Performance Computing; **PET**: Positron Emission Tomography; **CT**: Computed Tomography; **AR**: Augmented Reality; **GUI**: Graphical User Interface.

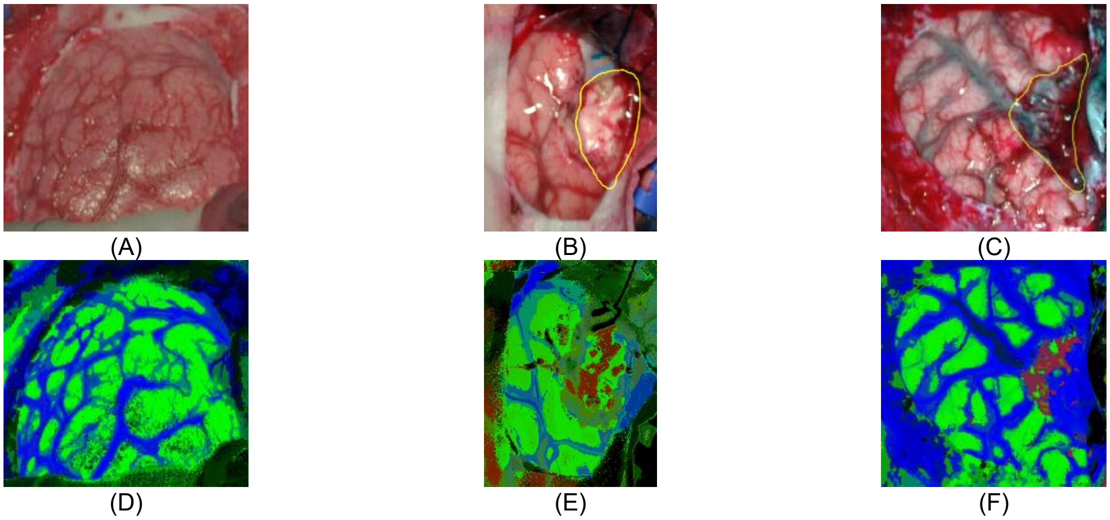

**Figure S2.** HELICoiD project classification results obtained from the validation database employed in [12]. (A, B, C) Synthetic RGB images; (D, E, F) Thematic maps of the HS image, where the tumour tissue is in red colour, the normal tissue in green, the hypervascularized tissue in blue and the background in black. (A, D) Normal brain tissue; (B, E) Primary grade 4 glioblastoma; (C, F) Primary grade 1 meningioma.

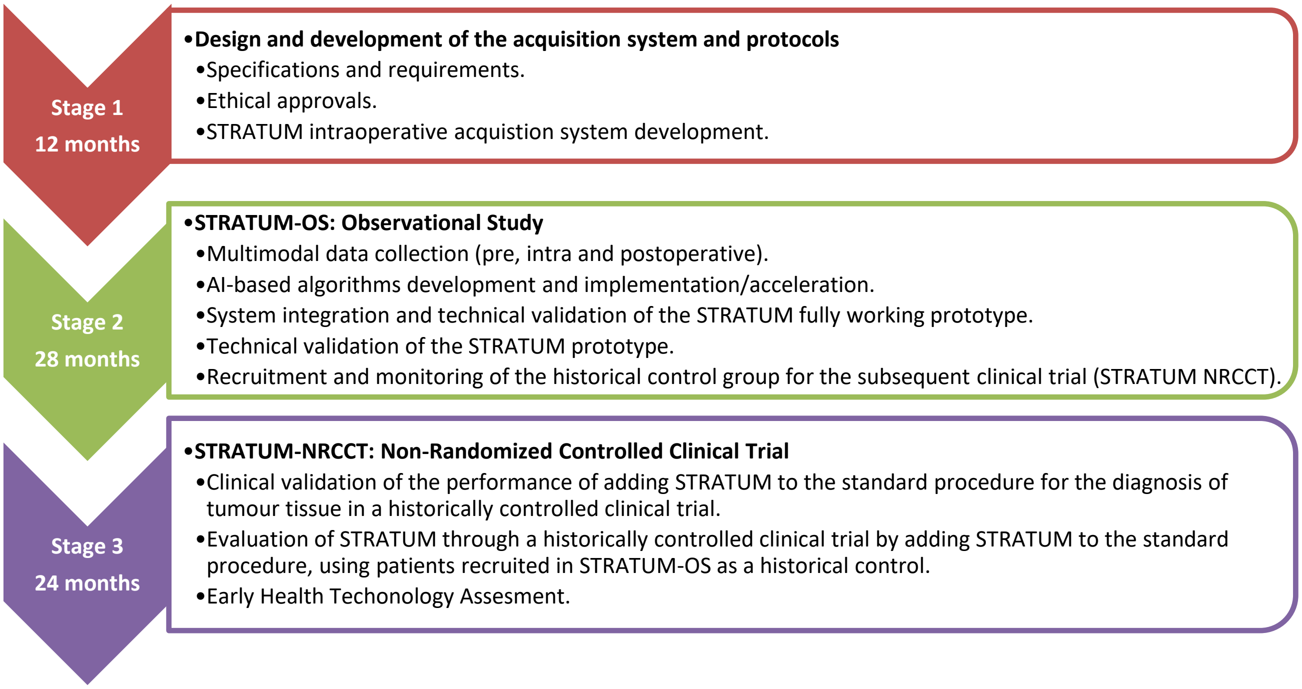

**Figure S3.** Work plan of the overall STRATUM project related to the two clinical studies that will be carried out in the project.

### SUPPLEMENTAL TABLES

**Table S1.** SPIROS 2023 Checklist: Recommended Items to address in the observational study Protocol and related documents**.** N/A: Not Applicable.

| Section / Item | Item Number | | Description | Page |
|---|---|---|---|---|
| **Part A: General information** | | | | |
| Title | 1 | □ | Descriptive title Identifying study design in the title | 1 (Title page) |
| Protocol version | 2 | □ | Version or amendment number with date and summary of the changes | 1 (Title page) |
| Protocol summary | 3 | □ | An informative and balanced summary of the study protocol | 2 |
| Sponsor and funder details | 4 | □ | Name of Sponsor and funder and types of financial, material, and other support | 9 |
| Conflict of interest statements | 5 | □ | Statement about any financial and other competing interests for principal or co-investigators for the overall study. | 9 |
| Investigators name | 6a | □ | Names of the principal and co-investigators | 1 (Title page) |
| Affiliation of investigators | 6b | □ | Affiliated institutions of the investigators | 1 (Title page) |
| Principal researcher/s contact detail | 6c | □ | Name, e-mail address, affiliation of principal researcher | 1 (Title page) |
| **Part B: Introduction** | | | | |
| Background of the study | 7a | □ | Description of research question and scientific background of the study | 2 |
| Review of prior research | 7b | □ | Summary of relevant existing research (published or unpublished) | 2 |
| Rationale of study | 7c | □ | Justification for conducting the study | 2 |
| Aim | 8a | □ | Broader aims and overall objective | 2 |
| Objective/s of the study | 8b | □ | Primary and secondary objective/s including any prespecified hypothesis (if applicable). | 2-3 |
| | 8c | □ | Specify whether the intention is to (a) estimate causal effects, (b) predict outcomes, or (c) simple description. | 2-3 |
| **Part C: Methods** | | | | |
| Study design | 9a | □ | Description of study design (case control, cross-sectional or cohort) and type of study (retrospective cohort study, Prospective cohort study, etc.) | 3 |
| Study setting | 9b | □ | Description of the study setting (e.g., community-based, hospital based) and detail of precise locations of the study sites. | 4 |
| Study schedule | 10a | □ | Description of the expected schedule of the study including relevant dates, expected periods of recruitment/survey, exposure, follow-up, and data collection. | 3 |
| | 10b | □ | Figure (Study schematic/flow-chart) or table describing expected time frame for each step including trainings, data collection, follow-up, analysis and reporting etc. | 3 |
| Sample size | 11 | □ | Estimation of minimum sample size required for the study with justifications including clinical and statistical assumptions supporting any sample size calculations. | 3 & 7-8 |
| Sampling procedure | 12 | □ | Detailed description of the sampling frame and sampling strategy (simple random, stratified random, cluster, systematic etc.) | 3 & 7 |
| **Participant selection** | | | | |
| Participant selection for cohort study | 13a | □ | Description of inclusion and exclusion criteria, and the source and methods of participant selection (exposed and unexposed). For matched cohort studies, give matching criteria and number of exposed and unexposed. | 3 |
| Participant selection for case-control study | 13b | □ | Description of inclusion and exclusion criteria, and the source and methods of case ascertainment and control selection. Give the rationale for the choice of cases and controls. Give diagnostic criteria for identifying cases (if applicable). For matched case-control studies, give matching criteria and the number of controls per case. | N/A |
| Participant selection for cross-sectional study | 13c | □ | Description of the inclusion and exclusion criteria, and the source and methods of participant selection. | N/A |
| Variables | 14a | □ | Detailed description of all important baseline and outcome variables to be analysed, exposures, predictors, potential confounders, and effect modifiers. Give diagnostic criteria, if applicable. | 5-8 |
| Data sources/measurement | 14b | □ | For each variable of interest, give sources of data and details of assessment /measurement methods. Describe comparability of assessment methods if there is more than one group. | 5-8 & Supplemental Tables |
| Data collection and management | 15a | □ | Plans for assessment and collection of outcomes, baseline, follow up and other study related data. | 4-5 |
| | 15b | □ | Description of data collection methods e.g., online survey, Household survey, paper based or electronic data capture, etc. | 4-5 |
| | 15c | □ | Any related processes to promote data quality during data collection (e.g., duplicate measurements, training of assessors, validation method) | 4-5 |
| | 15d | □ | Description of study instruments (e.g., questionnaires, data collection forms) along with their reliability and validity, if known. Reference to where data collection forms can be found, if not in the protocol. | 4-5 |
| | 15e | □ | Plans for data entry, coding, security, and storage, including any related processes to promote data quality (e.g., electronic data capture, double data entry; range checks for data values, random cross-checking of electronic data with the source documents). | 4-5 & 8-9 |

| | | | | |
|---|---|---|---|---|
| | 15f | □ | Reference to where details of data management procedures can be found, if not in the protocol. | N/A |
| Blinding procedure (if blinded study) | 16 | □ | Description of blinding procedure (if applicable) reporting Who will be blinded (e.g., investigator blinded for disease status when measuring exposure in case-control study) and methods to ensure blinding and unmasking of blinding if required. | N/A |
| Potential bias | 17 | □ | Description of any potential biases and plan to minimize those potential sources of biases. | 7-8 |
| Statistical analysis plan | 18 | □ | Detailed description of methods for analyzing and presenting primary/secondary outcomes and any additional analysis (e.g. analyses of subgroups and interactions, and sensitivity analyses). Give reference to where other details of the statistical analysis plan can be found, if not in the protocol. | 7-8 |
| Handling of missing data | 19 | □ | Detailed description of methods to handle missing data (e.g. multiple imputation). | 7 |
| Handling of withdrawals and lost to follow up | 20a | □ | Detailed description of the procedures to be followed when a participant ceases participation in the study prematurely or is lost to follow up | 5 |
| Replacements | 20b | □ | Plans and methods of the replacement or substitution of withdrawn participants. | N/A |
| Outcome | 21 | □ | Definition and description of all primary, secondary and other outcomes. | 5-6 |
| Data confidentiality statement | 22 | □ | A detailed description of process to ensure data confidentiality. | 8-9 |
| Follow up | 23 | □ | A detailed plan of follow up including schedule and methods (telephonic, house based, hospital based etc.) of follow up. | 5-6 |
| Plan of study monitoring | 24 | □ | Description of plan for study monitoring and whether the monitoring will be independent from investigators or sponsors. | 5 |
| Training of surveyors/data collectors | 25 | □ | Description of how investigators and surveyors will be trained to conduct the research activity. | 3 |
| Quality assurance | 26 | □ | Plan of quality assurance. back-checking data collection. | 5 |
| **Part D: Ethical consideration** | | | | |
| Ethical approval | 27a | □ | Plan for seeking ethics approval from ethics committees/institutional review boards. If known, give name of ethical committees. | 8-9 |
| | 27b | □ | If ethics approval will not be sought, give justification. | N/A |
| Consent and assent | 28a | □ | Description of who will obtain informed consent or assent from potential study participants or authorized surrogates, and how (e.g., written informed consent, verbal consent, video/audio recording of consent procedure etc.) | 3 |
| | 28b | □ | Give reason if consent or assent not sought. | N/A |
| | 28c | □ | Give reference to where informed consent forms and applicable translations plan can be found, if not in the protocol. | 3 |
| Risk/harm to participants | 29a | □ | A detailed description of potential risks or harms to study participants. | 8 |
| | 29b | □ | Plans for collecting, assessing, reporting, and managing any study procedures related adverse events (e.g. adverse events due to blood collection) and other unintended effects of study conduct (e.g. risk to breach confidential and sensitive information of participants) | N/A |
| | 29c | □ | Give a statement about whether data will be anonymous, pseudonymized, or can be directly linked to participants. | 4-5 |
| | 29d | □ | Description of any plan for giving Incentives to the participants | N/A |
| Adverse event and serious adverse event reporting | 30 | □ | Outline how adverse events and serious adverse events information will be collected and reported. | N/A |
| Involvement of patient/participant representatives in protocol development | 31 | □ | Patient and Public Involvement (PPI) statement including how patients or participants involved in the planning of the study. Give statement, if there is no plan to involve of patient/participants and public in designing or any phase of the study | 2 |
| **Part E. Reporting and dissemination** | | | | |
| Dissemination/ publication plan | 32a | □ | Plans for investigators and sponsor to communicate study results to ethical review boards, participants, key stake holders, the public, and other relevant groups. | 8-9 |
| | 32b | □ | Methods to communicate findings (e.g., via publication (open access or closed access), reporting in results databases, or other data-sharing arrangements), including any publication restrictions. | 9 |
| | 32c | □ | Define authorship eligibility guidelines (e.g., ICMJE recommendations) | 9 |
| **Part F: Others** | | | | |
| Whether Artificial Intelligence (AI) assisted technology was used in writing the protocol | 33 | □ | Disclose whether authors used artificial intelligence (AI)- assisted technologies in the production of protocol (e.g., chatbots) or there is planning to use artificial intelligence (AI)- assisted technologies in the production of manuscript or study reports. | 9 |
| | 34 | □ | Give the name of AI tools (such as ChatGPT). Include a statement if authors did or did not review and edited the content created by AI-assisted technologies | N/A |
| References | 35 | □ | A complete list of references cited in protocol. | 10-11 |
| Funding | 36 | □ | Source of any funding for the study and the role of the funders for the study | 9 |
| Open science | 37a | □ | **Registration of observational study:** Study identifier and registry name (e.g., open science framework, ClinicalTrials.gov, ICTRP or any other national or international study registry platform). If not yet registered, name of intended registry. | 10 |
| | 37b | □ | **Data sharing:** Plans, if any, for granting public access to the (1) full protocol and amendments, (2) participant-level data set, (3) Statistical analysis plan, (4) statistical codes and other study material (e.g., case report forms, study questionnaires and Informed consent forms). Give reference to where these documents can be found, if not included as annex in the protocol. | 9 |

**Table S2.** STRATUM-OS variables related to the patient characteristics.

| Variable | Categories/Interval | Units | Responsible (Department) |
|---|---|---|---|
| Patient ID (anonymized) | ID (e.g., S-K-001) | - | DC (NS) |
| Age | ≥18 | Years | DC (NS) |
| Year of birth | yyyy | - | DC (NS) |
| Weight | – | Kg | DC (NS) |
| Sex | - Female<br>- Male | - | DC (NS) |
| Hospital ID | - KUH<br>- HUGCDN<br>- HU12O | - | DC (NS) |
| Comorbidities | - Hypertension<br>- Diabetes<br>- Heart disease<br>- Coagulopathy<br>- Treatment with antiaggregant or anticoagulant<br>- Charlson comorbidity index<br>- Other (please, specify) | - | DC (NS) |
| Handedness | - Right<br>- Left | - | DC (NS) |
| Previous brain tumour surgery | - Yes, same site<br>- Yes, other site<br>- No | - | DC (NS) |
| Previous brain tumour surgery: Diagnosis date | yyyy-mm-dd | - | DC (NS) |
| Previous brain tumour surgery: Surgery date | yyyy-mm-dd | - | DC (NS) |
| Family History of Cancer or Primary CNS Tumours | - Yes<br>- No | - | DC (NS) |
| Brain tumour diagnosis date | yyyy-mm-dd | - | DC (NS) |
| Palliative surgery | -Yes<br>-No | - | DC (NS) |
| Life expectancy < 6 months | -Yes<br>-No<br>-Not available | - | DC (NS) |

**DC:** Data collector; **NS**: Neurosurgery.

**Table S3.** STRATUM-OS variables related to the patient's symptoms.

| Variable | Categories/Interval | Units | Responsible (Department) |
|---|---|---|---|
| Motor deficits | - Unilateral weakness<br>- Bilateral weakness<br>- Plegia<br>- Ataxia<br>- Spasticity<br>- Loss of complex movement execution<br>- Other (specify) | - | DC (NS) |
| Language deficits | - Dysarthria<br>- Apraxia<br>- Other (specify) | - | DC (NS) |

**DC:** Data collector; **NS**: Neurosurgery.

**Table S4.** STRATUM-OS variables related to the PROMs (Patient-Reported Outcome Measures).

| Variable | Categories/Interval | Units | Responsible (Department) |
|---|---|---|---|
| KPS score | 0 to 100 | % | DC (NS) |
| Who is completing this questionnaire? | - The patient<br>- A family member or caregiver (proxy)<br>- Healthcare professional<br>- Other (please specify) | - | DC (NS) |
| Date questionnaire completed | yyyy-mm-dd | - | DC (NS) |
| ECOG score | 0 to 5 | - | DC (NS) |
| Who is completing this questionnaire? | - The patient<br>- A family member or caregiver (proxy)<br>- Healthcare professional<br>- Other (please specify) | - | DC (NS) |
| Date questionnaire completed | yyyy-mm-dd | - | DC (NS) |
| EQ-5D-5L value | -0.31 to 1 (Sweden)<br>-0.416 to 1 (Spain) | - | DC (NS) |
| Who is completing this questionnaire? | - The patient<br>- A family member or caregiver (proxy)<br>- Healthcare professional<br>- Other (please specify) | - | DC (NS) |
| Date questionnaire completed | yyyy-mm-dd | - | DC (NS) |
| EORTC QLQ-BN20 score | 0 to 100 | - | DC (NS) |
| Who is completing this questionnaire? | - The patient<br>- A family member or caregiver (proxy)<br>- Healthcare professional<br>- Other (please specify) | - | DC (NS) |
| Date questionnaire completed | yyyy-mm-dd | - | DC (NS) |
| EORTC QLQ-C30 score | 0 to 100 | - | DC (NS) |
| Who is completing this questionnaire? | - The patient<br>- A family member or caregiver (proxy)<br>- Healthcare professional<br>- Other (please specify) | - | DC (NS) |
| Date questionnaire completed | yyyy-mm-dd | - | DC (NS) |

**DC:** Data collector; **NS**: Neurosurgery.

**Table S5.** STRATUM-OS variables related to the Vasari guide.

| Variable | Categories/Interval | Units | Responsible (Department) |
|---|---|---|---|
| Date of diagnostic MRI | yyyy-mm-dd | - | DC (NR) |
| F1-Tumour Location | - Frontal lobe<br>- Temporal lobe<br>- Parietal lobe<br>- Occipital lobe<br>- Insular<br>- Basal ganglia<br>- Thalamus<br>- Brainstem<br>- Cerebellum<br>- Corpus callosum | - | DC (NR) |
| F2-Side of tumour epicentre | - Right<br>- Centre/Bilateral<br>- Left | - | DC (NR) |
| F3-Eloquent brain | - No eloquent brain<br>- Speech motor<br>- Speech receptive<br>- Motor<br>- Vision | - | DC (NR) |
| F4-Enhancement quality | - None<br>- Minimal/Mild<br>- Marked/Avid | - | DC (NR) |
| F5-Proportion enhancing | 0 to 100 | % | DC (NR) |
| F6-Proportion nCET | 0 to 100 | % | DC (NR) |
| F7-Proportion necrosis | 0 to 100 | % | DC (NR) |
| F8-Cyst(s) | - Yes<br>- No | - | DC (NR) |
| F9-Multifocal ormulticentric | - Focal<br>- Multifocal<br>- Multicentric<br>- Gliomatosis | - | DC (NR) |
| F10-T1/FLAIR ratio | - Expansive<br>- Mixed<br>- Infiltrative | - | DC (NR) |
| F11-Thickness of enhancing margin | - Thin (<3 mm)<br>- Thick/nodular (>3 mm)<br>- Solid | - | DC (NR) |
| F12-Definition of the enhancing margin | - Well-defined<br>- Poorly-defined<br>- n/a | - | DC (NR) |
| F13-Definition of the non-enhancing margin | - Well-defined<br>- Poorly-defined<br>- n/a | - | DC (NR) |
| F14-Proportion of edema | 0 to 100 | % | DC (NR) |
| F16-Haemorrhage | - Yes<br>- No | - | DC (NR) |
| F17-Diffusion characteristics | - Facilitated<br>- Restricted<br>- Mixed<br>- Indeterminate | - | DC (NR) |
| F18-Pial invasion | - Yes<br>- No | - | DC (NR) |
| F19-Ependymal extension | - Yes<br>- No | - | DC (NR) |
| F20-Cortical involvement | - Yes<br>- No | - | DC (NR) |
| F21-Deep white matter invasion | - No<br>- Internal Capsule<br>- Brainstem<br>- Corpus Callosum | - | DC (NR) |
| F22-nCET crosses midline | - Yes<br>- No | - | DC (NR) |
| F23-CET crosses midline | - Yes<br>- No | - | DC (NR) |
| F24-Satellites | - Yes<br>- No | - | DC (NR) |
| F25-Calvarial remodelling | - Yes<br>- No | - | DC (NR) |

**DC:** Data collector; **NR**: Neuroradiology.

**Table S6.** STRATUM-OS variables related to the surgery details.

| Variable | Categories/Interval | Units | Responsible (Department) |
|---|---|---|---|
| Main surgeon ID | ID (e.g., 01) | - | DC (NS) |
| Main surgeon age at surgery date | - | years | DC (NS) |
| Main surgeon years of experience in this type of surgery at surgery date | - | years | DC (NS) |
| Assistant surgeon ID | ID (e.g., 01) | - | DC (NS) |
| Assistant surgeon age at surgery date | - | years | DC (NS) |
| Assistant surgeon years of experience in this type of surgery at surgery date | - | years | DC (NS) |
| Surgery date | yyyy-mm-dd | - - | DC (NS) |
| Surgical tools applied | - IONM<br>- Ultrasonic Aspirator<br>- 5-ALA microscope<br>- 5-ALA exoscope<br>- Neuronavigation<br>- Ultrasound probe<br>- Navigated ultrasound<br>- Other (specify) | - | DC (NS) |
| Fluorescence use | - Not used<br>- Gliolan<br>- Fluorescein<br>- Other (specify) | - | DC (NS) |
| Fluorescence dose | - | mg | DC (NS) |
| Anaesthesia type | - Propofol<br>- Remifentanil infusion<br>- Dexmedetomidine<br>- Sevoflurane<br>- Other (specify) | - | DC (AE) |
| Anaesthesia dose | - | ml | DC (AE) |
| Total operation time | - | min | DC (NS) |
| Total surgical time | - | min | DC (NS) |
| Involved professionals during surgery: Type | - Main neurosurgeon<br>- Assistant neurosurgeon<br>- Anaesthetists<br>- Pathologist<br>- Neurophysiologist<br>- Main nurse<br>- Assistant Nurse<br>- Other (specify) | - | DC |
| Involved professionals during surgery: Time | - | min | DC |
| Waiting time for STRATUM acquisition system use during surgery | - | min | DC |
| Intraoperative diagnosis time | - | min | DC |
| Waiting time for intraoperative diagnosis | - | min | DC |

**DC:** Data collector; **NS**: Neurosurgery; **AE**: Anaesthesiology.

**Table S7.** STRATUM-OS variables related to the intraoperative pathology.

| Variable | Categories/Interval | Units | Responsible (Department) |
|---|---|---|---|
| Sample ID | ID (e.g., 01) | - | DC (NP) |
| Tissue type | - Non-tumour<br>- Tumour | - | DC (NP) |
| Tumour group (if *Tumour*) | - Primary<br>- Secondary | - | DC (NP) |
| Tumour grade (if *Primary*) | - High<br>- Low | - | DC (NP) |

**DC:** Data collector; **NP**: Neuropathology.

**Table S8.** STRATUM-OS variables related to the definitive pathology.

| Variable | Categories/Interval | Units | Responsible (Department) |
|---|---|---|---|
| Total time definitive pathology analysis | – | days | DC (NP) |
| Specimen size | - | mm | DC (NP) |
| Tissue type | - Non-tumour<br>- Tumour | - | DC (NP) |
| Tumour group (if *Tumour*) | - Primary<br>- Secondary | - | DC (NP) |
| Tumour grade (if *Primary*) | - G1<br>- G2<br>- G3<br>- G4 | - | DC (NP) |
| Tumour origin (if *Secondary*) | - Lung<br>- Breast<br>- Kidney<br>- Colon<br>- Skin<br>- Other (please, specify) | - | DC (NP) |
| Tumour entity | Categories of intra-axial tumours specified in **Annex 1** according to WHO CNS 2021. | - | DC (NP) |
| IDH mutation | - Positive<br>- Negative<br>- Not Performed | - | DC (NP) |
| ATRX mutation | - Positive<br>- Negative<br>- Not Performed | - | DC (NP) |
| P53 mutation | - Positive<br>- Negative<br>- Not Performed | - | DC (NP) |
| 1p19q deletion | - Positive<br>- Negative<br>- Not Performed | - | DC (NP) |
| BRAF mutation | - Positive<br>- Negative<br>- Not Performed | - | DC (NP) |

**DC:** Data collector; **NP**: Neuropathology.

**Table S9.** STRATUM-OS variables related to the definitive pathology (STRATUM-related samples).

| Variable | Categories/Interval | Units | Responsible (Department) |
|---|---|---|---|
| Sample ID | ID (e.g., 01) | - | DC (NP) |
| Specimen size | - | mm | DC (NP) |
| Tissue type | - Non-tumour<br>- Tumour | - | DC (NP) |
| Tumour grade (if *Tumour* and *Primary* in Table S7) | - G1<br>- G2<br>- G3<br>- G4 | - | DC (NP) |

**DC:** Data collector; **NP**: Neuropathology.

**Table S10.** STRATUM-OS variables related to the exitus.

| Variable | Categories/Interval | Units | Responsible (Department) |
|---|---|---|---|
| Exitus cause | - Related<br>- Non-related | - | DC |
| Exitus date | yyyy-mm-dd | - | DC |

**DC:** Data collector; **Related**: Exitus caused by the brain tumour; **Non-related:** Exitus caused by other disease/cause.

**Table S11.** STRATUM-OS variables related to the postoperative MRI outcomes.

| Variable | Categories/Interval | Units | Responsible (Department) |
|---|---|---|---|
| Date of postoperative MRI | yyyy-mm-dd | - | DC (NR) |
| Extent of tumour resection | - Gross Total Resection (GTR)<br>- Near-Total Resection (NTR)<br>- Subtotal Resection (STR)<br>- Not Resected (NR) | - | DC (NR) |
| Volume of residual tumour on postoperative MRI 48/72 h (=1.5T) | 0 to 100<br>[ - ] | %<br>[$cm^3$] | DC (NR) |

**DC:** Data collector; **NR**: Neuroradiology.

**Table S12.** STRATUM-OS variables related to the postoperative complications.

| Variable | Categories/Interval | Units | Responsible (Department) |
|---|---|---|---|
| Postoperative complication type | - Infection<br>- Blood clots<br>- Chest and breathing problems<br>- Bleeding<br>- Wound problems (e.g., the surgical wound opening)<br>- Allergic reaction to medication or blood products<br>- Other (specify) | - | DC (NS) |
| Treatment | - Free text (specify) | - | DC (NS) |
| Diagnostic test | - Free text (specify) | - | DC (NS) |
| Start date | yyyy-mm-dd | - | DC (NS) |
| End date | yyyy-mm-dd | - | DC (NS) |

**DC:** Data collector; **NS**: Neurosurgery.

**Table S13.** STRATUM-OS variables related to the postoperative medication.

| Variable | Categories/Interval | Units | Responsible (Department) |
|---|---|---|---|
| Medication group type | - Painkillers<br>- Steroids<br>- Anti-seizures<br>- Other related to the brain surgery (specify) | - | DC |
| Prescription Date | yyyy-mm-dd | - | DC |

**DC:** Data collector.

**Table S14.** STRATUM-OS variables related to the emergency and hospital readmissions.

| Variable | Categories/Interval | Units | Responsible (Department) |
|---|---|---|---|
| Visit to emergency? | - Yes<br>- No | - | DC |
| Visits to emergency room:<br>Admission date and time | yyyy-mm-dd<br>(HH:MM) | - | DC |
| Visits to emergency room:<br>Discharge date and time | yyyy-mm-dd<br>(HH:MM) | - | DC |
| Need for hospital readmission? | - Yes<br>- No | - | DC |
| Hospital readmissions:<br>Admission date | yyyy-mm-dd | - | DC |
| Hospital readmissions:<br>Discharge date | yyyy-mm-dd | - | DC |
| Need for revision neurosurgery | - Yes<br>- No | - | DC (NS) |

**DC:** Data collector; **NS**: Neurosurgery.

**Table S15.** STRATUM-OS variables related to the follow-up MRI.

| Variable | Categories/Interval | Units | Responsible (Department) |
|---|---|---|---|
| Date of follow-up MRI | yyyy-mm-dd | - | DC (NR) |
| MRI Reason | - Pre-treatment<br>- During treatment<br>- Post-treatment<br>- Recurrence<br>- Other (specify) | - | DC (NR) |
| Progression | - Yes<br>- No | - | DC (NR) |

**DC:** Data collector; **NR**: Neuroradiology.

**Table S16.** STRATUM-OS variables related to the postoperative treatments.

| Variable | Categories/Interval | Units | Responsible (Department) |
|---|---|---|---|
| Post-operative treatment class | - Chemotherapy<br>- Radiotherapy | - | DC (OC) |
| Post-operative treatment type (if *Chemotherapy*) | - Temozolomide<br>- Procarbazine<br>- Carmustine (BCNU)<br>- Lomustine (CCNU)<br>- Vincristine<br>- A combination of drugs called PCV (procarbazine, lomustine and vincristine)<br>- Other related (specify) | - | DC (OC) |
| Post-operative treatment type (if *Radiotherapy*) | - External beam radiation therapy (EBRT)<br>- Internal radiation (brachytherapy)<br>- Other related (specify) | - | DC (OC) |
| Post-operative treatment sub-type (if *Radiotherapy based on EBRT*) | - 3-D conformal therapy (3D-CRT)<br>- Intensity modulated radiation therapy (IMRT)<br>- Conformal proton beam therapy<br>- Stereotactic radiosurgery (SRS) based on gamma knife radiation<br>- Linear accelerator based SRS<br>- Other related (specify) | - | DC (OC) |
| #Cycles (if *Chemotherapy*) | – | #Cycles | DC (OC) |
| Dose (if *Chemotherapy*) | – | mg | DC (OC) |
| #Sessions (if *Radiotherapy*) | – | #Sessions | DC (OC) |
| Start date | yyyy-mm-dd | - | DC (OC) |
| End date | yyyy-mm-dd | - | DC (OC) |

**DC:** Data collector; **OC**: Oncology.

**Table S17.** STRATUM-OS variables related to the hospital stay of the patient.

| Variable | Categories/Interval | Units | Responsible (Department) |
|---|---|---|---|
| Stay at neurosurgical care ward: Admission date and time | yyyy-mm-dd (HH:MM) | - | DC |
| Stay at neurosurgical care ward: Discharge date and time | yyyy-mm-dd (HH:MM) | - | DC |
| Stay at hospital: Admission date | yyyy-mm-dd | - | DC |
| Stay at hospital: Discharge date | yyyy-mm-dd | - | DC |

**DC:** Data collector.

**Table S18.** STRATUM-OS variables related to the postoperative follow-up visits to professionals.

| Variable | Categories/Interval | Units | Responsible (Department) |
|---|---|---|---|
| Follow-up visits to professionals: Type | - Neurosurgeon<br>- Neurologist<br>- Oncologist<br>- Physiotherapists<br>- Speech and language therapists<br>- Occupational therapists<br>- Other (specify) | - | DC |
| Follow-up visits to professionals: Date | yyyy-mm-dd | - | DC |

**DC:** Data collector.

**Table S19.** STRATUM-OS variables related to the postoperative follow-up tests.

| Variable | Categories/Interval | Units | Responsible (Department) |
|---|---|---|---|
| Follow-up test date | yyyy-mm-dd | - | DC |
| Follow-up test type | - Blood test (related to follow-up brain surgery)<br>- Brain MRI scan<br>- Brain CT scan<br>- Other (specify) | - | DC |
| Blood test result (if *Blood test* type) | - Haemoglobin (g/dL)<br>- Haematocrits (%)<br>- Platelets (103/µL)<br>- Neutrophils (103/µL)<br>- Creatinine (mg/dL) | - | DC |

**DC:** Data collector.

**Table S20.** STRATUM-OS primary and secondary outcomes for technical validation.

| Variable | Categories/Interval | Units | Responsible (Department) |
|---|---|---|---|
| Sensitivity in tumour identification against anatomopathological analysis | 0 to 100 | % | DC (ST) |
| Specificity in tumour identification against anatomopathological analysis | 0 to 100 | % | DC (ST) |
| Sensitivity in identification of CET | 0 to 100 | % | DC (ST) |
| Specificity in identification of CET | 0 to 100 | % | DC (ST) |
| Sensitivity in identification of nCET (FLAIR positive) | 0 to 100 | % | DC (ST) |
| Specificity in identification of nCET (FLAIR positive) | 0 to 100 | % | DC (ST) |

**CET**: Contrast Enhanced Tumour; **nCET**: non-Contrast Enhanced Tumour; **DC**: Data Collector; **ST**: Statistics.

#### ANNEX 1: LIST OF INTRA-AXIAL TUMOUR ENTITIES ACCORDING TO THE INTEGRATED DIAGNOSIS (CNS WHO 2021)

___ Gliomas, glioneuronal tumors, and neuronal tumors

*Adult-type diffuse gliomas*

**___ Astrocytoma, IDH-mutant**
**___ Oligodendroglioma, IDH-mutant and 1p/19q-codeleted**
**___ Glioblastoma, IDH-wildtype**

*Pediatric-type diffuse low-grade gliomas*

**___ Diffuse astrocytoma, MYB- or MYBL1-altered**
**___ Angiocentric glioma**
**___ Polymorphous low-grade neuroepithelial tumor of the young**
**___ Diffuse low-grade glioma, MAPK pathway-altered**

*Pediatric-type diffuse high-grade gliomas*

**___ Diffuse midline glioma, H3 K27-altered**
**___ Diffuse hemispheric glioma, H3 G34-mutant**
**___ Diffuse pediatric-type high-grade glioma, H3-wildtype and IDH-wildtype**
**___ Infant-type hemispheric glioma**

*Circumscribed astrocytic gliomas*

**___ Pilocytic astrocytoma**
**___ High-grade astrocytoma with piloid features**
**___ Pleomorphic xanthoastrocytoma**
**___ Subependymal giant cell astrocytoma**
**___ Chordoid glioma**
**___ Astroblastoma, MN1-altered**

*Glioneuronal and neuronal tumors*

**___ Ganglioglioma**
**___ Gangliocytoma**
**___ Desmoplastic infantile ganglioglioma**
**___ Desmoplastic infantile astrocytoma**
**___ Dysembryoplastic neuroepithelial tumor**
**___ Diffuse glioneuronal tumor with oligodendroglioma-like features and nuclear clusters**
**___ Papillary glioneuronal tumor**
**___ Rosette-forming glioneuronal tumor**
**___ Myxoid glioneuronal tumor**
**___ Diffuse leptomeningeal glioneuronal tumor**
**___ Multinodular and vacuolating neuronal tumor**
**___ Dysplastic cerebellar gangliocytoma (Lhermitte-Duclos disease)**
**___ Central neurocytoma**
**___ Extraventricular neurocytoma**
**___ Cerebellar liponeurocytoma**

*Ependymal tumors*

**___ Supratentorial ependymoma**
**___ Supratentorial ependymoma, ZFTA fusion-positive**
**___ Supratentorial ependymoma, YAP1 fusion-positive**
**___ Posterior fossa ependymoma**
**___ Posterior fossa group A (PFA) ependymoma**
**___ Posterior fossa group B (PFB) ependymoma**
**___ Spinal ependymoma**
**___ Spinal ependymoma, MYCN-amplified**
**___ Myxopapillary ependymoma**
**___ Subependymoma**

___ Choroid plexus tumors

___ **Choroid plexus papilloma**
___ **Atypical choroid plexus papilloma**
___ **Choroid plexus carcinoma**

___ Medulloblastoma
*Must select both molecularly defined and histologically defined subtypes*
*Molecularly Defined Medulloblastomas*
___ **Medulloblastoma, WNT-activated**
___ **Medulloblastoma, SHH-activated and TP53-wildtype**
___ **Medulloblastoma, SHH-activated and TP53-mutant**
___ **Medulloblastoma, non-WNT / non-SHH**
*Histologically Defined Medulloblastomas*
___ **Classic medulloblastoma**
___ **Desmoplastic / nodular medulloblastoma**
___ **Medulloblastoma with extensive nodularity**
___ **Large cell / anaplastic medulloblastoma**

___ Other CNS embryonal tumors
___ **Atypical teratoid / rhabdoid tumor**
___ **Cribriform neuroepithelial tumor**
___ **Embryonal tumor with multilayered rosettes**
___ **CNS neuroblastoma, FOXR2-activated**
___ **CNS tumor with BCOR internal tandem duplication**
___ **CNS embryonal tumor, NEC / NOS**

___ Mesenchymal, non-meningothelial tumors involving the CNS
*Vascular tumors*
___ **Hemangioma**
___ **Cavernous malformation**
___ **Arteriovenous malformation**
___ **Capillary telangiectasia**
___ **Hemangioblastoma**

___ Hematolymphoid tumors involving the CNS
*CNS Lymphomas*
___ **Primary diffuse large B-cell lymphoma of the CNS**
___ **Immunodeficiency-associated CNS lymphoma**
___ **Lymphomatoid granulomatosis**
___ **Intravascular large B-cell lymphoma**
*Miscellaneous rare lymphomas in the CNS*
___ **MALT lymphoma of the dura**
___ **Other low-grade B-cell lymphomas of the CNS (specify subtype(s), if known): ___**
___ **Anaplastic large cell lymphoma (ALK+ / ALK-)**
___ **T-cell lymphoma**
___ **NK / T-cell lymphoma**

___ Germ cell tumors
___ **Mature teratoma**
___ **Immature teratoma**
___ **Teratoma with somatic-type malignancy**
___ **Germinoma**
___ **Embryonal carcinoma**
___ **Yolk sac tumor**

___ **Choriocarcinoma**
___ **Mixed germ cell tumor (specify subtype(s), if known): ________________**

___ Other (e.g., NEC, NOS) (specify): ________________
